# 2024/25 end-of-season KP.2 vaccine effectiveness against COVID-19 hospitalization in older adults: a test-negative study in Quebec, Canada

**DOI:** 10.64898/2026.04.02.26350050

**Authors:** Sara Carazo, Danuta M Skowronski, Chantal Sauvageau, Denis Talbot, Etienne Racine, Nicholas Brousseau

**Author notes:** Corresponding author: Sara Carazo, Institut national de santé publique du Québec, 945, Avenue Wolfe, Québec, Québec, G1V 5B3, Canada.

## Abstract

We evaluated 2024/25 KP.2 vaccine effectiveness (VE) against COVID-19 hospitalization among adults ≥60 years old eligible for publicly-funded vaccination during fall and/or spring campaigns in the province of Quebec, Canada. We included Quebec residents tested for COVID-19-compatible symptoms in an acute-care hospital between October 13, 2024 (epi-week 2024-42) and August 23, 2025 (2025-34), linking vaccine, hospital, chronic diseases and laboratory administrative records to assess VE through test-negative design. We compared the odds of being COVID-19 test-positive versus test-negative among vaccinated versus non-vaccinated participants, adjusting for sex, age, comorbidities, place of residence, and epidemiological week. Overall, 49,949 (43%) participants were vaccinated. Over an analysis period spanning up to ten months, including median time since vaccination of 16 weeks (interquartile range 9-24 weeks), VE was 34% overall, declining from 43% <8 weeks to negligible by the 32nd week post-vaccination. Findings confirm meaningful but short-lived COVID-19 vaccine protection against hospitalization in older adults.

**Highlights:**

- We estimate 2024/25 KP.2 vaccine effectiveness (VE) among adults aged ≥60 years
- Vaccination reduced the COVID-19 hospitalization risk by one-third overall
- Spring vaccine uptake was low, with shorter follow-up but VE like the fall analysis
- VE waned from about 40% at <8 weeks to negligible at ≥32 weeks post-vaccination
- COVID-19 VE against severe outcomes is meaningful but short-lived in older adults

## INTRODUCTION

In the province of Quebec, Canada, the updated KP.2 vaccine was recommended for the 2024/25 season, with the primary objective of reducing COVID-19-related hospitalizations and deaths [1]. During fall 2024, the Quebec immunization program targeted adults aged 60 years and older and individuals 6 months and older with high-risk conditions associated with a higher likelihood of severe outcomes. In contrast, the spring campaign focused on adults aged 75 years and older and immunocompromised individuals of any age [2]. Other non-prioritized individuals aged 6 months and older could also request a free COVID-19 vaccine dose during the 2024/25 season.

Early-season vaccine effectiveness (VE) estimates up to four months post-vaccination indicated moderate protection against COVID-19 hospitalization [3,4], while studies with longer follow-up past the fifth to seven months post-vaccination showed indication of waning effectiveness [5–7]. Overall, however, evidence on the durability of protection beyond six months remains limited [8], and no VE data are yet available for the 2025 spring vaccination campaign. This ten-month evaluation of the 2024/25 COVID-19 vaccination campaign presents KP.2 VE and duration of protection among older adults residing in Quebec, including both fall and spring estimates.

## METHODS

This test-negative study included all Quebec adults 60 years and older who were tested for COVID-19-compatible symptoms in an acute-care hospital between October 13, 2024 (epi-week 2024-42) and August 23, 2025 (2025-34).

Information on subvariant circulation was obtained from provincial SARS-CoV-2 genomic surveillance until March 2025, as described elsewhere [9], and from Canadian sources thereafter [10,11], when Quebec surveillance was suspended. In summary, KP.3 was predominant in October until mid-November 2024, followed by cocirculation of KP.3, XEC and LF.7 from December 2024 to March 2025 and by cocirculation of LP.8.1, NB.1.8.1 and XFG from April to August 2025. The lack of individual genotyping and a single predominant subvariant precluded subvariant-specific VE estimation for the 2024/25 season.

As previously described [9], we linked provincial vaccine, hospital, chronic disease surveillance and laboratory administrative databases to obtain population-level data on vaccination status, COVID-19 testing and hospitalizations.

We excluded tests conducted within 90 days following a positive test (predefined period to identify reinfections), negative tests performed within 7 days prior to a positive test (potential false negatives) or within 14 days after a negative test (assumed to be the same respiratory episode). We also excluded tests from individuals vaccinated <7 days before specimen collection, who had invalid vaccination dates or intervals, who were immunocompromised, or who resided in long-term care facilities (due to different hospitalization practices).

The fall vaccination campaign against COVID-19, influenza and respiratory syncytial viruses began on October 6, 2024, while the COVID-19-only spring campaign started April 6, 2025. Vaccination was defined as the receipt of ≥1 KP.2 vaccination at least 7 days before specimen collection and administered during the 2024/25 season (epi-weeks 2024-40 to 2025-34). Fall vaccination was from 2024-40 to 2025-15, and spring vaccination from 2025-16 to 2025-34 (Figure 1). Individuals tested during the spring vaccination and vaccinated only with a fall dose were excluded from the spring-specific analysis. Time since vaccination was evaluated using a categorical variable defined by eight-week incremental spans (e.g., ∼2-month intervals) since vaccination. Individuals were considered unvaccinated if they had received no COVID-19 vaccine or a non-KP.2 vaccine ≥6 months before specimen collection. Specimens were excluded if KP.2 vaccinated <7 days earlier or non-KP.2 vaccinated <6 months earlier.

**Figure 1.**
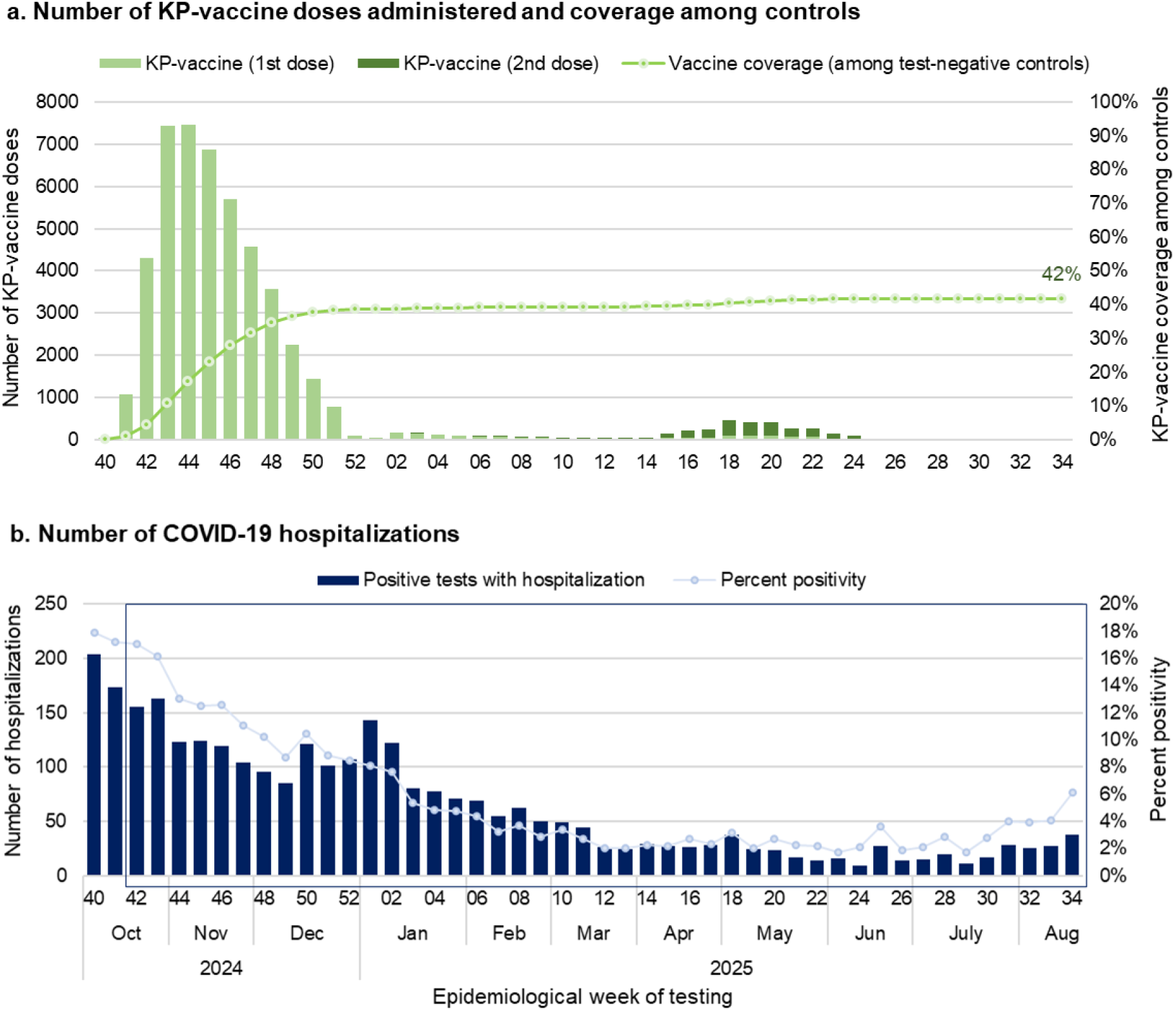
KP.2 vaccine administration and COVID-19 hospitalizations among adults 60 years and older in the province of Quebec. Note: The blue box in the second panel represents the study period

Cases were defined as SARS-CoV-2 positive tests performed in an acute-care hospital for COVID-19 compatible symptoms and followed (0-7 days later) by a hospital admission of ≥24 hours with a COVID-19 diagnosis as reason for admission, while negative tests (whether followed or not by hospitalization) were controls.

Logistic regression models compared the odds of KP.2 vaccination between cases and controls adjusting for sex, age, comorbidities, place of residence (home or private homes for older people) and epi-week. VE was calculated as (1-adjusted odds ratio (OR))*100 with 95% confidence intervals (95%CI). VE was estimated globally for the entire 2024/25 season and separately for the fall and spring campaigns, by time since vaccination, by number of KP.2 doses and by six-week calendar periods. The same covariables were used in stratified analyses, but time was adjusted using two-week intervals.

This study was conducted under the legal mandate of the Quebec National Director of Public Health and was approved by the ethics committee of the *Centre de Recherche du CHU de Québec-Université Laval (2021-5773)*.

## RESULTS

Among 157,670 SARS-CoV-2 tests performed in Quebec acute-care hospitals, 41,603 (26.4%) were excluded. The main reasons for exclusion included: tests involving immunocompromised individuals (n=10,054; 6.4%); positive tests without subsequent hospitalization (n=3,958; 2.5%); tests performed within 90 days of a previous positive result (3,995; 2.5%); and negative tests performed within 14 days of a negative test (14,748; 9.4%) (Figure 2).

**Figure 2.**
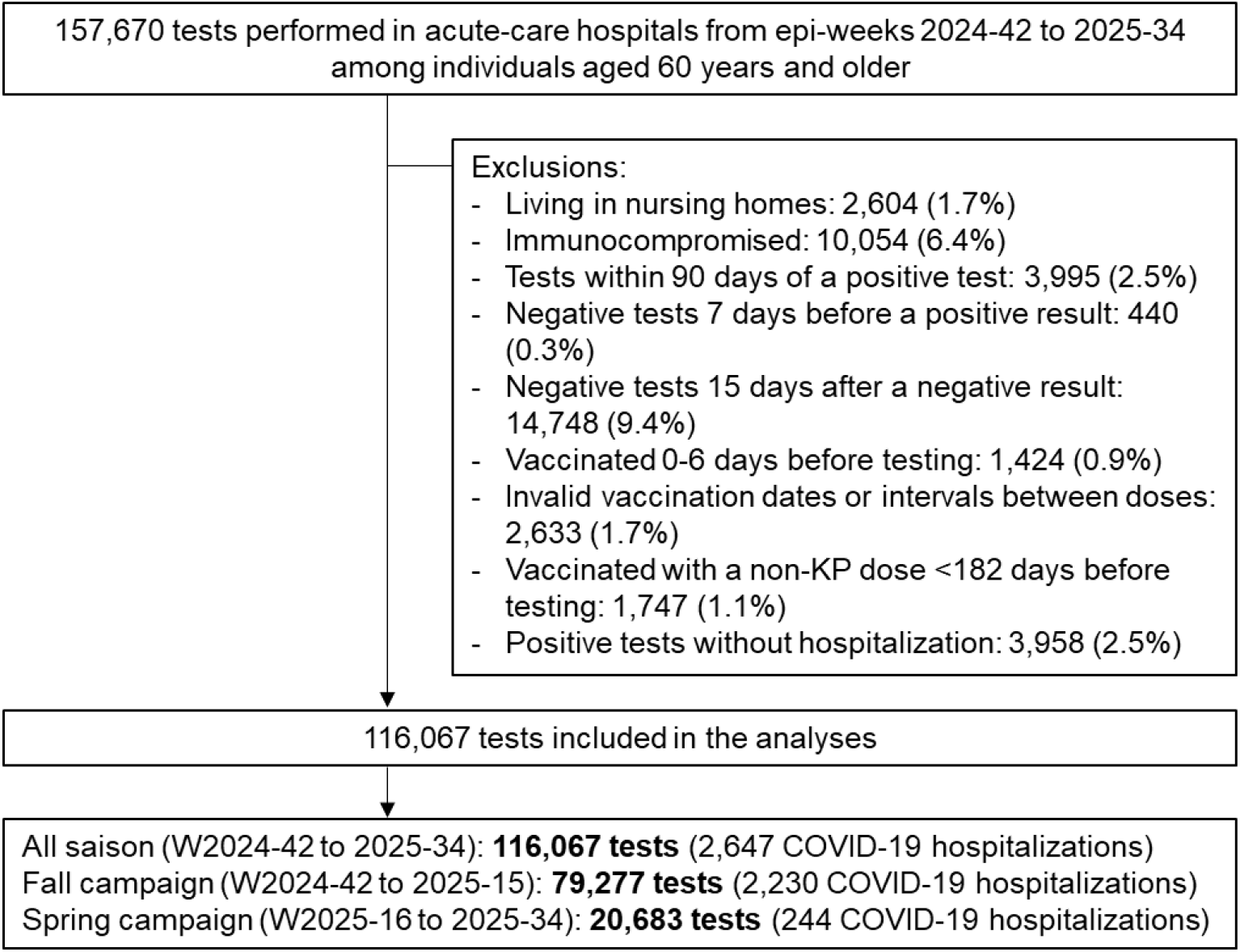
Study flowchart. Abbreviations: w, epi-week Note: 15,451 tests conducted in epi-weeks 2025-16 to 2025-34 among individuals vaccinated during the fall campaign were not included in the specific spring analyses

Overall, 49,949 (43.0%) participants (29.6% of cases and 43.3% of controls) were KP.2 vaccinated; 18,768 (38.2%) with BNT162b2 (Pfizer-BioNTech) and 30,398 (61.8%) with mRNA-1273 (Moderna) (Table 1). Among test-negative controls, 46,602 (41.1%) received a fall dose, increasing by age group 60-69, 70-79 and ≥80 years old at 27.5%, 41.4%, and 51.2%, respectively, each similar if slightly lower than reported provincial vaccine coverages for these age groups within the general population at 30%, 48%, and 56%, respectively [12]. A spring dose was administered to just 2,564 (2.3%) of controls, with only 1.8% receiving both doses (Figure 1, Table 1).

**Table 1.**
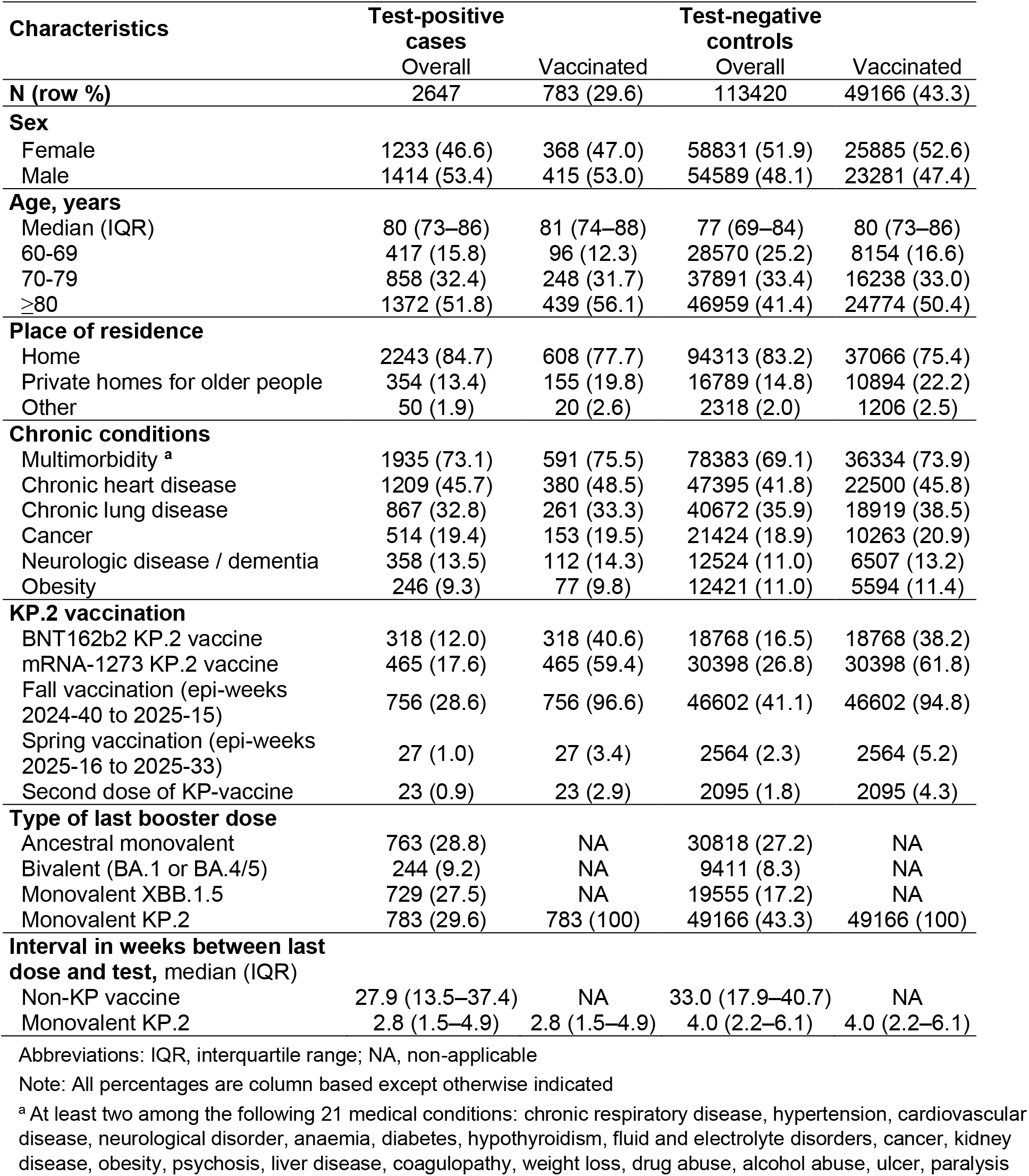
Characteristics of study participants.

From a peak of 155 hospitalized cases and 18% test-positivity in epi-week 2024-42, COVID-19 decreased steadily over the study period with between 9 and 69 cases and <5% positivity between epi-weeks 2025-06 and 2025-32 (Figure 1). Overall, 2,644 hospitalizations contributed to the full season analysis, including 2,230 in the fall analysis and 248 in the spring analysis (Figure 2).

Global KP.2 VE for the 2024/25 season was 34% (95%CI:28-40) at a median time since vaccination of 16 weeks (IQR=9-24) (Figure 3). One- and two-dose KP.2 VE estimates were similar at 34% (95%CI:28-40) and 35% (95%CI:-4-60), respectively. Fall VE was estimated at 38% (95%CI:31-34) at a median time since vaccination of 12 weeks and spring VE at 38% (95%CI:3-60) at a median time since vaccination of 7 weeks. Spring VE remained unchanged (VE=38%, 95%CI:0-61) when restricted to the age group eligible for vaccination (individuals ≥75 years old). Younger participants had lower and more imprecise VE estimates than older groups (Figure 3).

**Figure 3.**
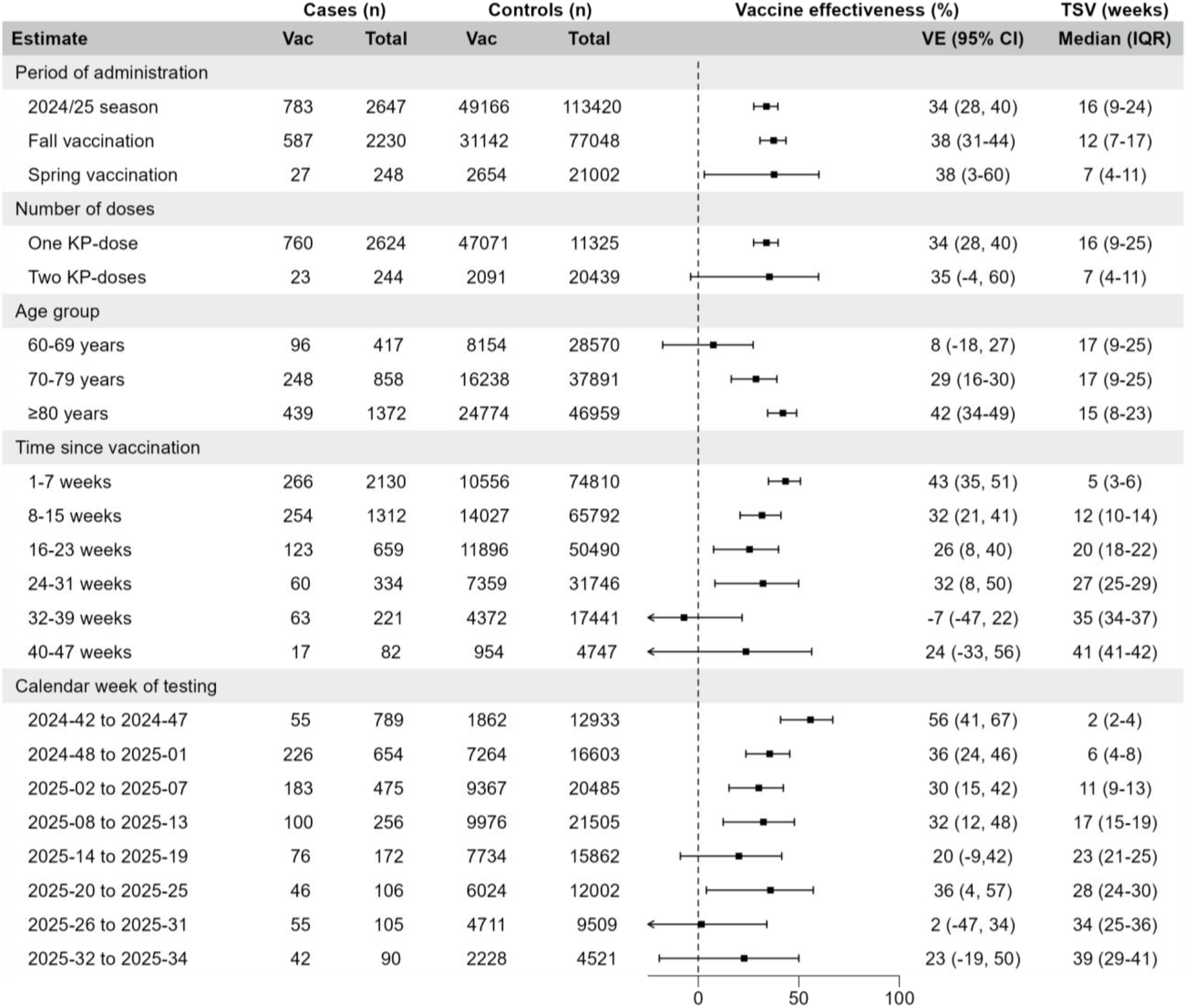
KP.2 vaccine effectiveness against COVID-19 hospitalization (epi-weeks 2024-42 to 2025-34) Abbreviations: CI, confidence interval; IQR, interquartile range; TSV, time since vaccination; Vac, vaccinated; VE, vaccine effectiveness. Note 1: Logistic regression model adjusted for sex, age group, multimorbidity, chronic respiratory disease, chronic cardiac disease, obesity, cancer, neurological disease, place of residence and epi-week (2-week periods for the time since vaccination analyses) compared KP.2 vaccinated participants with participants who did not receive the KP.2 vaccine. Note 2: Study period (epi-weeks) for each estimate was defined as follows: 2024/25 season (2024-42 to 2025-34), fall vaccination (2024-42 to 2025-15), spring vaccination (2025-16 to 2025-34), one KP-dose (2024-42 to 2025-34), two KP-doses (2025-16 to 2025-34), age groups ((2024-42 to 2025-34), TSV of: 1-7 weeks (2024-42 to 2025-34), 8-15 weeks (2024-49 to 2025-34), 16-23 weeks (2025-05 to 2025-34), 24-31 weeks (2025-13 to 2025-34), 32-39 weeks (2025-21 to 2025-34), 40-47 weeks (2025-30 to 2025-34), calendar week of testing (epi-weeks as indicated).

VE waned with time since vaccination, from 43% at <8 weeks since vaccination to ∼30% at 8 to 31 weeks and negligible after 32 weeks post-vaccination for a total follow-up of 47 weeks (∼10 months) (Figure 2). VE estimates also decreased with calendar time when examined by six-week periods (Figure 2).

## DISCUSSION

In this end-of-season population-based analysis, we showed that the updated 2024/25 mRNA KP.2 vaccine reduced the risk of COVID-19 hospitalization by about 40% in older adults during the first two months post-vaccination, but that vaccine protection waned mid-season, becoming negligible by the seventh month post-vaccination.

In the early post-pandemic era, yearly updated vaccines attempt to address the continuous genomic evolution of the SARS-CoV-2 virus. These variant-targeted vaccines are marketed with limited antigenicity or immunogenicity and no efficacy data, requiring real-world seasonal evaluation of their observed effectiveness.

Early 2024/25 season estimates from the USA reported KP.2 VE against hospitalizations of 45% at a median time since vaccination of two months [3], while European studies, where the JN.1 vaccine was administered, reported higher VE of 60-70% [4,13]. End-of-season studies, mostly conducted until April 2025, found global KP.2 VE estimates against hospitalization ranging from 20% to 40%, which aligns with our fall estimate of 38% for the October-April period [5,7,8,14]. Most studies reported a decline in effectiveness over time since KP.2-vaccination with a follow-up extending up to four or five months [5–8,15]. In our study, which spans a period of ten months, we observed an early decline in VE from 43% (7-60 days post-vaccination) to 30% (61-182 days), with no measurable protection beyond 6 months (183-304 days). A comparable absence of protection (VE=26%, 95%CI:-31-61) was also noted in the only (preprint) study to report effectiveness estimates beyond 180 days post-vaccination [8].

Evidence from previous seasons from Canada and elsewhere showed that initial moderate protection progressively declined over the season, potentially due to both waning effectiveness and variant replacement [9,16,17]. In the 2024/25 season, no differential protection has been reported against XEC vs non-XEC (mainly KP.3.1.1 and descendants) in studies evaluating KP.2 [8,11] or JN.1 vaccines [13]. However, European countries using vaccines targeting JN.1 lineage reported higher VE than USA or Canada, suggesting that JN.1 vaccines may have offered superior protection than KP.2-adapted vaccines against SARS-CoV-2 lineages circulating during the 2024-25 season [4,5,7,8,13]. LP.8.1 emerged late during the season, and its contribution was not captured in most studies. Although we did not have individual data on SARS-CoV-2 lineages in our study, our low end-of season VE estimates, as well as the reported LP.8.1 lineage-specific low VE against hospitalization (VE=24%, 95%CI:-19-53) [8] could be attributed to both lineage immune evasion and time since vaccination.

The absence of clear SARS-CoV-2 seasonality to date, together with evidence of low to negligible protection beyond 6 months post-vaccination, has prompted some countries, like Canada, UK and Australia, to recommend a spring dose to high-risk groups [6,9,11,17,18]. To address unpredictable seasonality, year-round protection seems necessary since SARS-CoV-2 circulation can peak in the summer, right before the fall vaccination campaign, as was the case during the 2024-25 season [19]. No other study to date has reported the 2025 spring VE which we estimated at 38%. Though less precise due to low uptake, spring VE was similar to the fall VE at shorter time since vaccination (7 versus 12 weeks) suggesting a possible decrease in protection against lineages circulating at the end of the season. We note, however, that the spring VE of 38% at median 7 weeks also does not differ meaningfully from the fall VE of 43% at 7-60 days (median 5 weeks) post-vaccination.

Our study has several limitations. We relied on administrative databases and although we used an algorithm incorporating test results and reasons for admission to identify hospitalizations attributable to COVID-19, some misclassification remains possible. Since SARS-CoV-2 circulation peaked just before the fall campaign, infected individuals might have postponed vaccination. Infection-acquired immunity among unvaccinated participants may introduce a downward bias in VE estimates. However, self-awareness of SARS-CoV-2 infection is low due to limited testing outside hospital settings. We could not evaluate the relative impact of waning protection versus lineage replacement for two reasons: (1) we did not have information on individual virus genotyping, and (2) the 2024/25 season was characterized by mixed cocirculation of multiple subvariants without a single dominating genotype. We observed an unexpected lower VE among the younger age group (60-69 years old), which had the lowest vaccine coverage. This might be partially explained by chance variation and/or residual confounding if younger vaccinated individuals have unmeasured characteristics associated with COVID-19 hospitalization risk. We could not adjust for influenza vaccination or restrict analyses to non-influenza controls, as this information was not available. However, the potential confounding arising from correlation between COVID-19 and influenza vaccination, both recommended in the fall campaign, is expected to be minimal, based on findings from studies assessing COVID-19 VE during the 2024-25 season [11,20].

Our results, together with the available evidence, reinforce impressions of meaningful yet short-lived protection of fall COVID-19 vaccination against hospitalization. We also evaluated the spring campaign and showed similar effectiveness, but the impact of this campaign was also limited by very low viral circulation and vaccine uptake. Our findings as well as the overall decline in COVID-19 burden within the general population [3,9] underscores the importance of prioritizing high-risk individuals for routinely repeated vaccination and research toward more effective and durable vaccines.

## Author contributions

All authors had full access to all of the data in the study and take responsibility for the integrity of the data and the accuracy of the data analysis. **Concept and design**: SC. **Acquisition, analysis, or interpretation of data**: All authors. **Drafting of the manuscript**: SC. **Critical revision of the manuscript for important intellectual content**: NB, CS, ER, DT, DMS. **Statistical analysis**: SC.

## Data availability

Underlying data cannot be shared publicly by authors because it belongs to the Ministère de la santé et des services sociaux du Québec and data access to researchers was given under the legal mandate of the Quebec National Director of Public Health.

